# Novel Privacy Considerations for Large Scale Proteomics

**DOI:** 10.1101/2022.04.06.22269907

**Authors:** Andrew C. Hill, Elizabeth M. Litkowski, Ani Manichaikul, Leslie Lange, Katherine A. Pratte, Katerina J. Kechris, Matthew DeCamp, Marilyn Coors, Victor E. Ortega, Stephen S. Rich, Jerome I. Rotter, Robert E. Gerzsten, Clary B. Clish, Jeffery Curtis, Xiaowei Hu, Debby Ngo, Wanda K O’Neal, Deborah Meyers, Eugene Bleecker, Brian D. Hobbs, Michael H. Cho, Farnoush Banaeikashani, Russell P. Bowler

## Abstract

**Introduction:** Privacy protection is a core principle of genomic research but needs further refinement for high-throughput proteomic platforms.

**Methods:** We identified independent single nucleotide polymorphism (SNP) quantitative trait loci (pQTL) from COPDGene and Jackson Heart Study (JHS) and then calculated genotype probabilities by protein level for each protein-genotype combination (training). Using the most significant 100 proteins, we applied a naïve Bayesian approach to match proteomes to genomes for 2,812 independent subjects from COPDGene, JHS, SubPopulations and InteRmediate Outcome Measures In COPD Study (SPIROMICS) and Multi-Ethnic Study of Atherosclerosis (MESA) with SomaScan 1.3K proteomes and also 2,646 COPDGene subjects with SomaScan 5K proteomes (testing). We tested whether subtracting mean genotype effect for each pQTL SNP would obscure genetic identity.

**Results:** In the four testing cohorts, we were able to correctly match 90%-95% their proteomes to their correct genome and for 95%-99% we could match the proteome to the 1% most likely genome. With larger profiling (SomaScan 5K), correct identification was > 99%. The accuracy of matching in subjects with African ancestry was lower (∼60%) unless training included diverse subjects. Mean genotype effect adjustment reduced identification accuracy nearly to random guess.

**Conclusion:** Large proteomic datasets (> 1,000 proteins) can be accurately linked to a specific genome through pQTL knowledge and should not be considered deidentified. These findings suggest that large scale proteomic data be given privacy protections of genomic data, or that bioinformatic transformations (such as adjustment for genotype effect) should be applied to obfuscate identity.

## Introduction

Nearly four decades ago Jeffreys et al [1] recognized that patterns of simple tandem-repetitive regions of DNA were specific for individuals and could be used for identifying specific individuals or close relatives. Although initially controversial, the DNA-fingerprinting technique was rapidly and widely adapted by forensic scientists and within a decade was in the public’s vernacular. Soon thereafter the results of the Human Genome Project were published [2, 3] and it is now recognized that there are millions of single nucleotide polymorphisms (SNP) which can distinguish individuals within large populations. Identifying individuals by genomics is a rising concern in research because advances in genotyping and sequencing have resulted in large genetic databases (dbGaP; GEO; EMBL-EBI) for both research and commercial use. The existence of newer genotyping technologies and large genomic databases has created concerns among policy makers regarding discrimination in health insurance and employment and resulted in new laws that address genetic information (e.g., the Genetic Information Non-discrimination Act of 2008) as well as privacy protection efforts such as the Global Alliance for Genomics and Health, which has created frameworks to ensure responsible and secure sharing of genomic and health-related data. A key feature of these policies in the United States is that they explicitly addressed genomic (single nucleotide, sequence, transcriptome, epigenomic, and gene expression) data only. Despite these policies, there have been multiple instances of “deidentified” personal information linked back to individual genetic profiles [4], including well publicized individuals such as Henrietta Lacks [5]. There have also been methods proposed which can link expression data to genotype through eQTLs [6].

Although lagging behind genotype and sequencing advances by 5-10 years, exponential technological advances in high throughput proteomics are leading to the creation of similar large databases with sensitive personal information. Concurrently there are studies which demonstrate that many proteins [7, 8] have genetic quantitative trait loci (QTLs), but current practice is to consider these datasets as deidentified data. In this manuscript we show that even limited proteome profiles without peptide sequencing can be linked to specific individuals by using prior independent knowledge of these QTLs and we provide a bioinformatic solution which obfuscates reidentification, yet still preserves at least some biomarker-phenotype relationships. These findings suggest an immediate need to change policy regarding non-genomic data used for research or commercial use.

## Methods

### Study Populations

All study participants provided written informed consent approved by institutional review boards (IRBs). COPDGene and Jackson Heart Study (JHS) cohorts were randomly split into training and testing datasets and training subjects were not included in the testing cohort. Other independent cohorts used for testing included Subpopulations and Intermediate Outcome Measures in COPD Study (SPIROMICS) and Multi-Ethnic Study of Atherosclerosis (MESA). Race was self-reported. Characteristics of subjects used for training and test are shown below with summary demographics in **Table 1**.

**Table 1:**
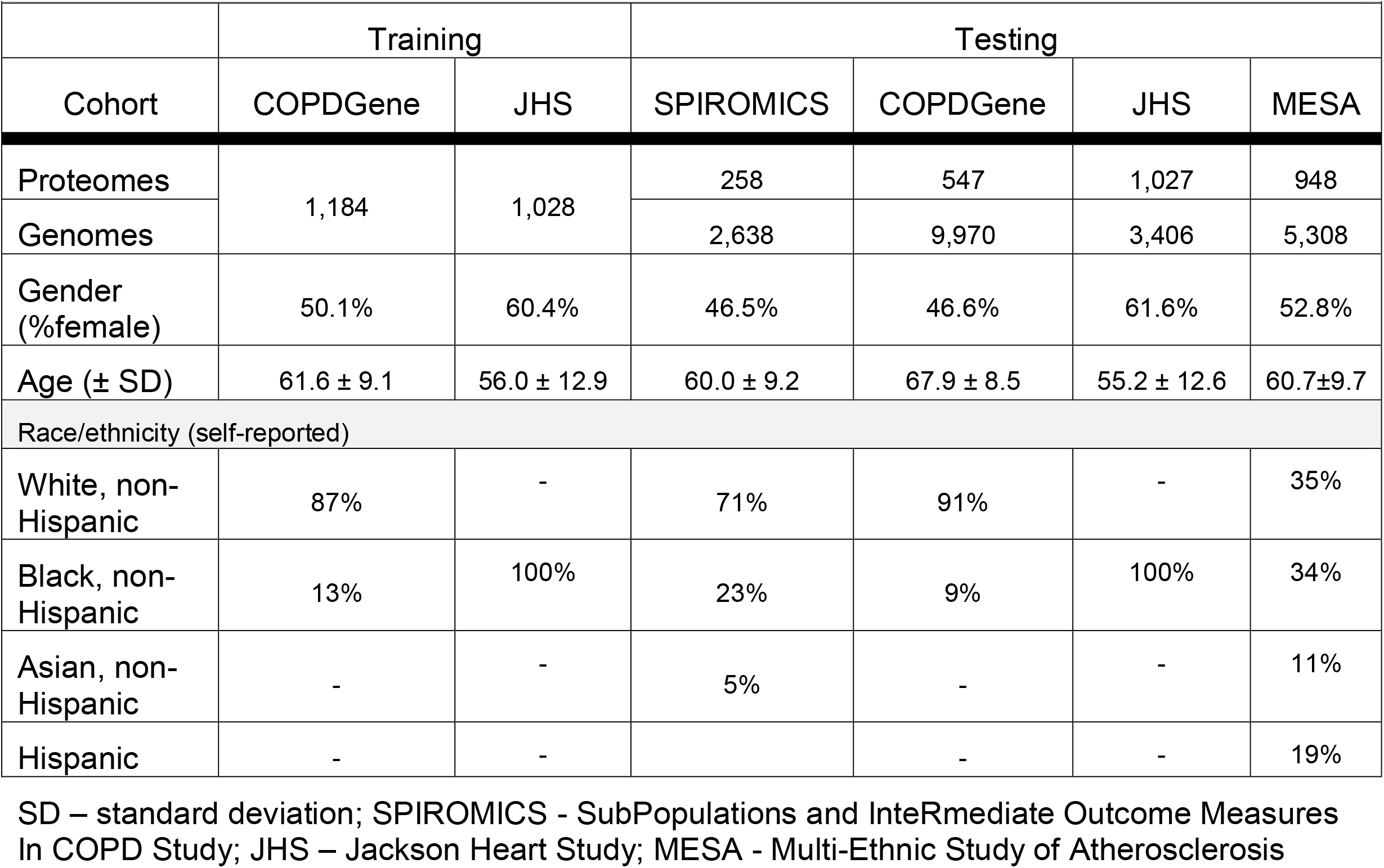
Characteristics of training cohort and independent testing cohorts with SomaScan 1.3K

|  |  |  |  |  |  |  |
| --- | --- | --- | --- | --- | --- | --- |
|  | Training |  | Testing |  |  |  |
| Cohort | COPDGene | JHS | SPIROMICS | COPDGene | JHS | MESA |
| Proteomes | 1,184 | 1,028 | 258 | 547 | 1,027 | 948 |
| Genomes |  |  | 2,638 | 9,970 | 3,406 | 5,308 |
| Gender (%female) | 50.1% | 60.4% | 46.5% | 46.6% | 61.6% | 52.8% |
| Age (± SD) | 61.6 ± 9.1 | 56.0 ± 12.9 | 60.0 ± 9.2 | 67.9 ± 8.5 | 55.2 ± 12.6 | 60.7±9.7 |
| Race/ethnicity (self-reported) |  |  |  |  |  |  |
| White, non-Hispanic | 87% | - | 71% | 91% | - | 35% |
| Black, non-Hispanic | 13% | 100% | 23% | 9% | 100% | 34% |
| Asian, non-Hispanic | - | - | 5% | - | - | 11% |
| Hispanic | - | - |  | - | - | 19% |

SD – standard deviation; SPIROMICS - SubPopulations and InteRmediate Outcome Measures In COPD Study; JHS – Jackson Heart Study; MESA - Multi-Ethnic Study of Atherosclerosis

#### COPDGene

The NIH-sponsored multicenter Genetic Epidemiology of COPD (COPDGene (ClinicalTrials.gov Identifier: NCT01969344)) enrolled 10,263 non-Hispanic white (NHW) and Black (AA) individuals from January 2008 until April 2011 (Phase 1) who were aged 45-80 with ≥10 pack-year smoking history and no exacerbations for >30 days and 457 age and gender matched healthy individuals with no history of smoking were enrolled as controls [9]. Subjects were genotyped using an Illumina HumanOmni Express [10]. 1,184 subjects from the enrollment visit (P1) participated in an ancillary study in which they provided p100 (BD) fresh frozen plasma used for SomaScan 1.3K proteomic profiling which measured 1,305 proteins.

An additional 547 independent subjects, who only had SomaScan profiling at a 5-year follow up visit (P2) and not used in the training dataset, were used as an independent testing cohort. 5,292 also had SomaScan 5k (v4.0) proteomes using plasma from a P2 visit and were randomly split into training and testing to assess whether scaling improved identification accuracy. *Data sets and availability*. Genotype data and SomaScan can be found on dbGaP for COPDGene (<u>phs000179.v6.p2</u>) and include “CG10k_NHW_hg19_Oct2017{.bed,.bim,.fam}” for NHW Subjects and “CG10k_AA_hg19_Oct2017{.bed,.bim,.fam}” for AA Subjects [7].

#### Jackson Heart Study (JHS)

The NIH-sponsored (ClinicalTrials.gov Identifier: NCT00005485) enrolled 5,306 African American residents living in the Jackson, MS, metropolitan statistical area (MSA) of Hinds, Madison, and Rankin Counties. 2,055 gave consent for genetic research and also had SomaScan 1.3K proteomic profiling. Genotypes were extracted using TOPMed whole genome sequencing Freeze 8 to create a synthetic Illumina HumanOmniExpress genotype panel. *Data sets and availability*. Genotype data can be requested through TOPMed and SomaScan can be found on dbGaP (phs000964).

#### SPIROMICS

The NIH-sponsored Subpopulations and Intermediate Outcome Measures in COPD study (SPIROMICS) study (ClinicalTrials.gov Identifier: NCT01969344) [11] enrolled 2,984 subjects who were genotyped using the Illumina HumanOmniExpress genotyping platform [12] of which 258 subjects underwent SomaScan 1.3K proteomic profiling using Visit 1 plasma. *Data sets and availability*. SPIROMICS dataset used include “SPIRO_SUBJID_2638{.bed,.bim,.fam}. Genotype data and SomaScan can be found on dbGaP (phs18817).

#### MESA

The NIH-sponsored Multi-Ethnic Study of Atherosclerosis (MESA) study (ClinicalTrials.gov Identifier: NCT00005487) recruited 6,418 participants from four race/ethnic groups: Caucasian, African American, Hispanic, and Chinese. Whole genome sequencing (WGS) was performed at the Broad Institute of MIT and Harvard. SomaScan proteomics 1.3K profiling was performed at the Broad Institute and Beth Israel Proteomics Platform (HHSN268201600034I). *Data sets and availability*. Genotype and SomaScan data can be requested through TOPMed and dbGaP (phs001416.v1.p1).

### Proteome Profiling

Proteomic profiles for 1,305 proteins were generated using SomaScan v 1.3K (SomaLogic, Boulder, Colorado). Description of the SomaScan 1.3k assay is further described in [13]. Normalization follow SomaLogic’s guidelines for data processing encompass three sequential levels of normalization, namely Hybridization Control Normalization (Hyb) followed by Median Signal Normalization (Hyb.MedNorm) and Interplate Calibration (Hyb.MedNorm.Cal). There are no missing data on the platform. SomaScan 5K v4.0 (4,776 proteins) was performed by SomaLogic and we used Adaptive Normalization by Maximum Likelihood (anmlSMP). For pQTL discovery, we used a rank-based inverse normal transformation to align protein levels to a normal distribution; however, for estimating genotype probabilities and associations with smoking, we used log transformed protein values.

### Statistical analyses

#### pQTL discovery by protein wide association study (pWAS)

COPDGene had genotyping for 691,764 SNPs without imputation. Genotype for these SNPs in JHS were called using TOPMed whole genome sequence. Only SNPs with minor allele frequencies (MAF) greater than 5% in the sample population were included for analysis. Both datasets were aligned to GRCh38. SNP-by-proteins associations were assessed in separately in both the COPDGene and JHS discovery cohorts using linear regression assuming an additive model by genotype. Analysis was performed using the R package ‘MatrixEQTL’ (version 2.2) [14]. Each model assessed direct association between protein level and genotype, with no adjustment for covariates. Protein quantitative trait loci (pQTLs) were considered significant at FDR corrected p-value < 0.05. The pQTL assessments in JHS and COPDGene were performed independently. After merging the two sets of pQTLs from the two training cohorts, we reduced the set to obtain a list of uniquely associated protein and SNP combinations. For each unique protein in the pQTL set, we kept only the highest significance SNP pQTL as determined by the p-value for the training cohorts (**Figure 1**). When the two training cohorts had different top SNPs (often in linkage disequilibrium), we chose the SNP from the cohort with the lowest p-value. This first-level reduction produces a set of unique proteins, but in some cases, multiple proteins may be associated with the same SNP. If a SNP was associated with multiple proteins, we used only the protein with the highest protein association for that SNP. This process ensured that each protein and each SNP appear only once in our pQTL sets.

**Figure 1.**
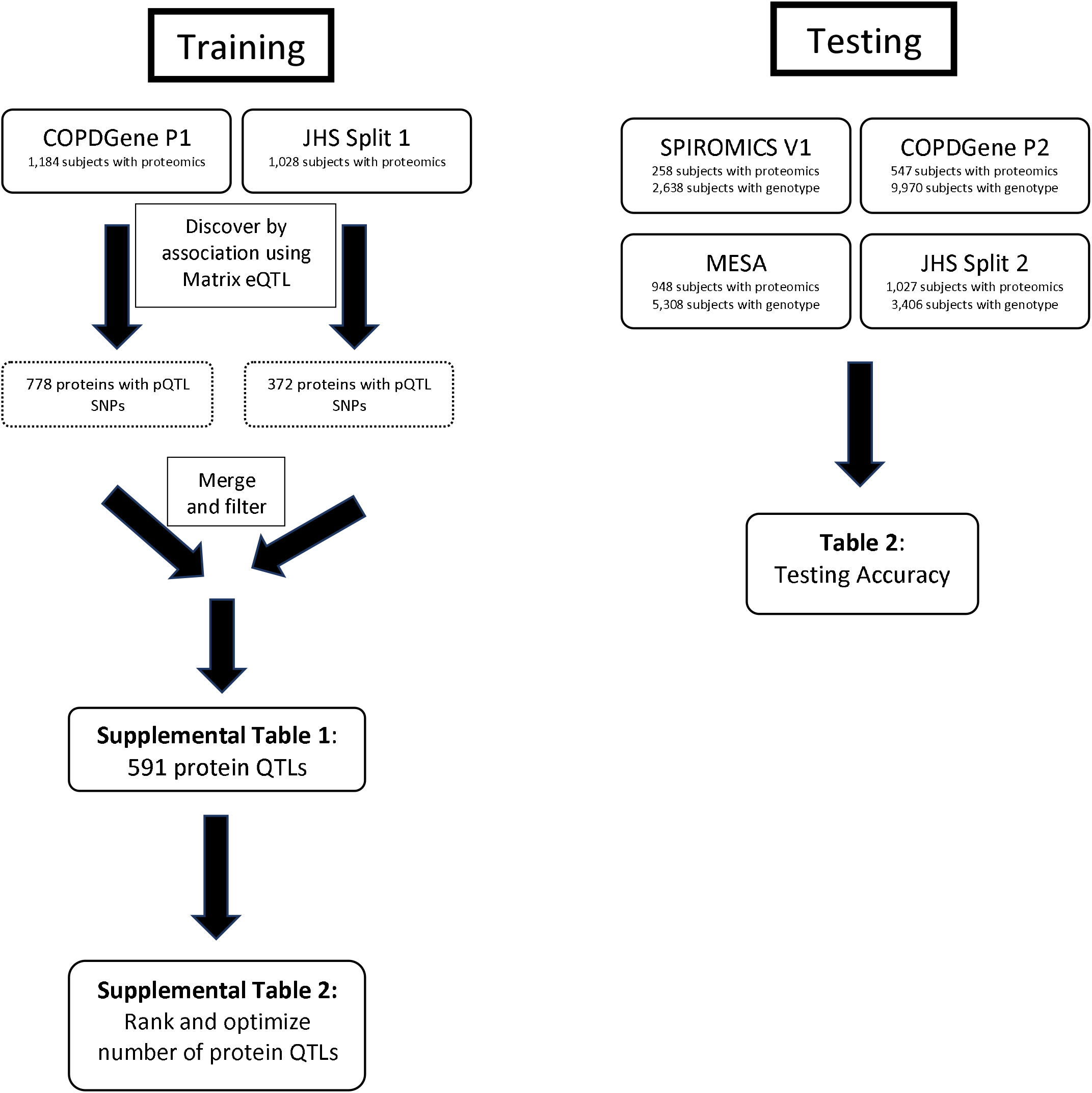
Strategy for identifying protein-QTL SNP combinations (training) and testing accuracy of proteins for identifying the subject by association with genotypes file.

#### Bayesian modeling

For predicting the probability of a genome matching, we use a Naïve Bayesian method (**Figure 2**) which estimates the probability of observing genotype vector *g* using the genotype specific mean (µ) and standard deviation (σ) estimated from training data. This is similar to an approach used in genotype estimation from eQTLs [6]. To combine the training estimates from COPDGene and JHS we used the GaussianNB model from scikit-learn (version 0.23.2) for this estimation [6]. During training, we use the partial_fit method to calculate µ and σ parameters on a single dataset. The same method can be used to update parameters µ and σ, allowing us to train a model on multiple datasets by sharing the trained model. Since each SNP is biallelic, we calculate three probabilities corresponding to the three possible genotypes.

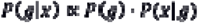

using a Gaussian naïve Bayes framework, where we define three normal probability distribution functions

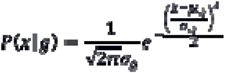

which describe the distribution of protein levels for each of the three genotypes (**Figure 3a**), where μ_g_ and σ_g_ are the estimated mean and variance respectively of the protein levels *x* for subjects with genotype *g*. Under the naïve Bayes framework, we estimate the probability of the subject possessing each of the three genotype classes, given an observed protein level (**Figure 3b**). By repeating this process for each of the protein/SNP pairs, we obtain the probability of each genotype class for the top 100 SNPs. We calculate the odds of each genotype being the true genotype, and then using the known genotype values for each subject, we can compute the odds of observing the correct or “true” genotype vector for a subject as the product of the odds of observing the individual true genotype values.

**Figure 2.**
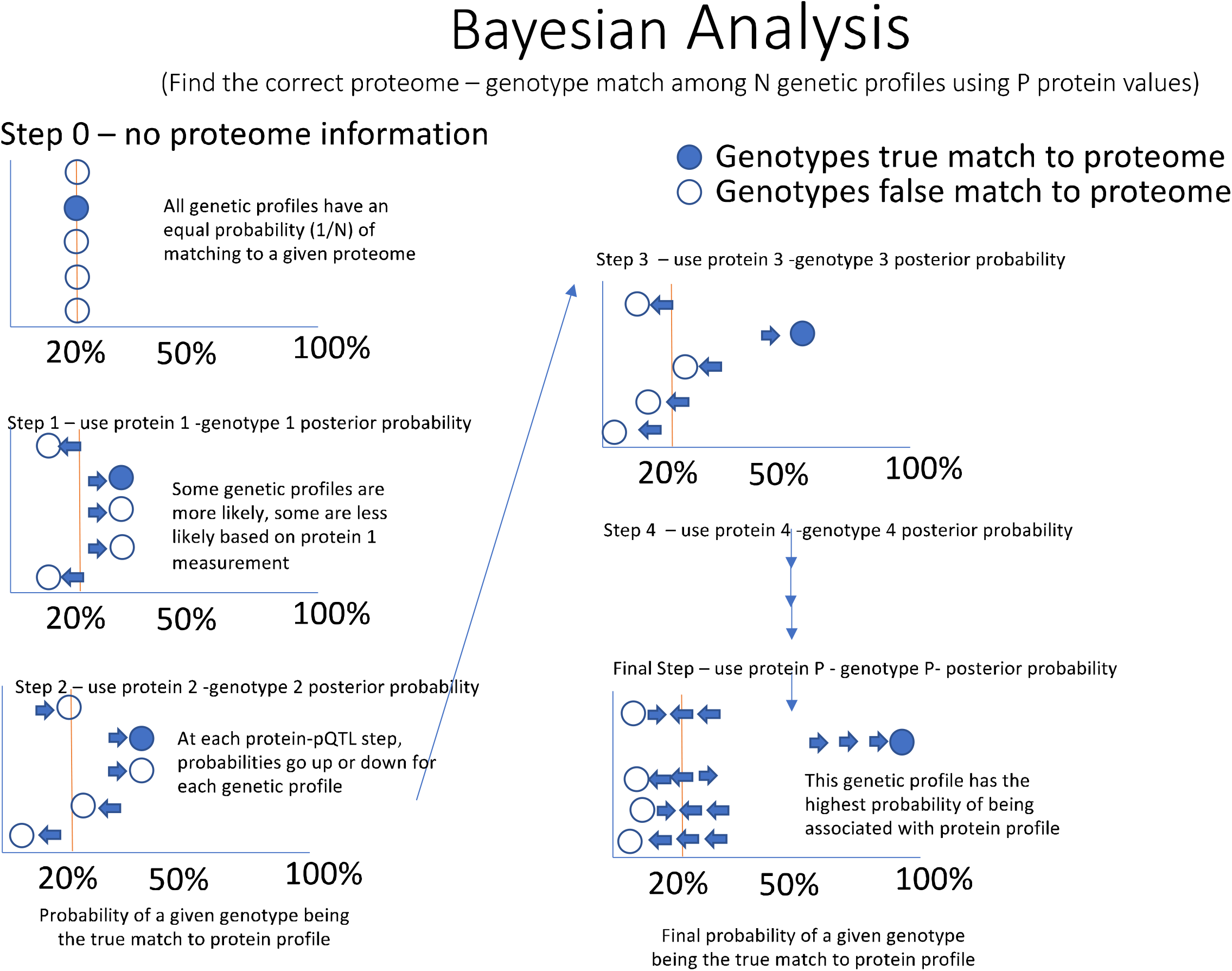
Naïve Bayes approach to estimate posterior probability of a subject matching genotype predicted by protein levels

**Figure 3.**
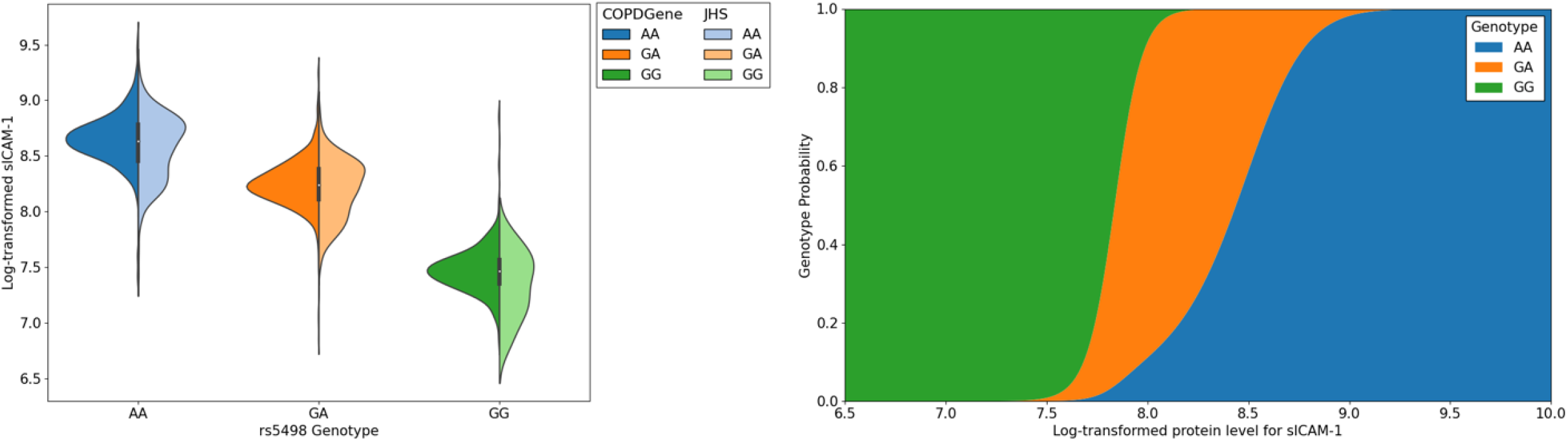
(**A**). Beeswarm showing the protein distributions for sICAM-1, which have been log transformed and stratified by genotype in COPDGene and JHS training sets. In this example AA is the major genotype. (**B**). Probability function for genotype by protein value for sICAM-1.

For each subject with proteome data, we calculate the odds of the genotype vector of every genotyped subject in the dataset. Assuming one of the genotyped subjects within the dataset is the true identity with observed protein levels we take the genotype with the highest odds given the observed protein values as the “match” for this subject. If the genotype with the highest odds of match (top 1) belongs to the subject whose protein levels were observed, we consider this a match. We also tested whether the true match was among the three highest odds (top 3) and 1% highest odds (in top 1%).

#### Associations with smoking

A T-test was used to assess whether proteins (log transformed) were associated with current smoking (smoking cigarettes in the past 30 days).

#### Software and packages

All analyses were run in R (version 3.6.11) and Python (version 3.7)

## Results

### Model training and parameter optimization

Our first training attempts at model training used only COPDGene subjects, which were mostly subjects with predominant European ancestry. This analysis identified 778 proteins with at least one pQTL SNP. To test the accuracy of protein measurements to predict genotypes, every proteome was assigned a probability of proteome matching genome (**Figure 4**). The accuracy of the method was determined by how many times a subject with a proteome had the true genome assigned the highest probability of a match as the first choice, top three choices, or top 1% of the dataset. This method demonstrated excellent testing accuracy in identifying independent subjects of European ancestry in COPDGene, MESA, and SPIROMICS (83-92%); however, testing accuracy in subjects with predominantly African ancestry was significantly lower (61%-76%) (**Table 2**). Therefore, we retrained our models using additional African-Ancestry subjects from JHS subjects. In the JHS training data set we identified 372 proteins with at least one pQTL SNP. We then combined the COPDGene and JHS training pQTLs for a total of 591 proteins with at least one pQTL SNP (**Supplemental File 1**). Using these combined COPDGene and JHS training set we significantly improved the matching accuracy in African American subjects (**Figure 5**) which improved accuracy to ∼90%, which is similar to accuracy in European ancestry subjects.

**Table 2.** Accuracy of matching proteome profiles to genetic profiles using 150 proteins **s**

|  |  |  |  |  |
| --- | --- | --- | --- | --- |
|  |  | % correctly identified |  |  |
| Testing Cohort | Subgroup | Top 1 | In top 3 | In Top 1 % |
| COPDGene | Overall | 85.0% | 89.0% | 97.8% |
|  | NHW | 86.0% | 89.6% | 98.4% |
|  | AFA | 75.5% | 83.7% | 91.8% |
| JHS | AFA | 85.8% | 91.5% | 98.1% |
| MESA | NHW | 92.4% | 96.1% | 98.3% |
|  | AFA | 61.0% | 73.6% | 86.8% |
|  | Chinese-American | 85.5% | 94.2% | 100% |
|  | Hispanic | 88.9% | 93.8% | 96.9% |
| SPIROMICS | Overall | 93.4% | 98.5% | 99.2% |
|  | NHW | 92.4% | 97.8% | 98.9% |
|  | AFA | 96.7% | 100% | 100% |
|  | Other | 92.9% | 100% | 100% |

**Figure 4.**
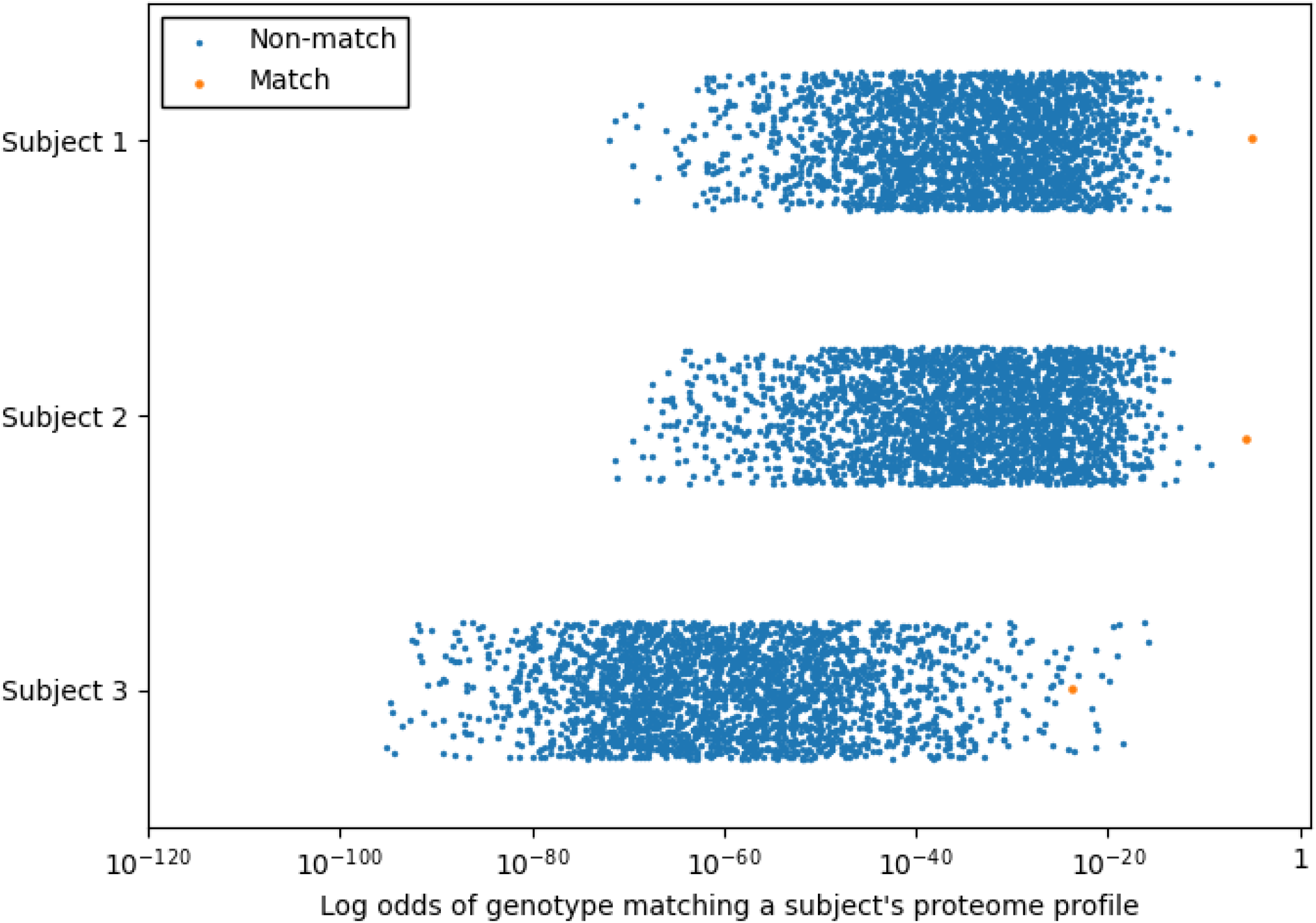
Probability that a proteome matches a given genome in the test dataset. In this example, 100 proteins are used to identify probable genotype at 100 pQTL SNPs. The majority of proteome profiles were associated with the correct genotype profile (orange circle) with near 100% probability of being correct (Subject 1 and 2). The rest of the proteome profiles typically were represented in the top 1% of highest probability genotypes matches (top 26 of 2,698) as demonstrated by Subject 3. The blue circles probability of genotype profile matching from incorrect subjects.

**Figure 5.**
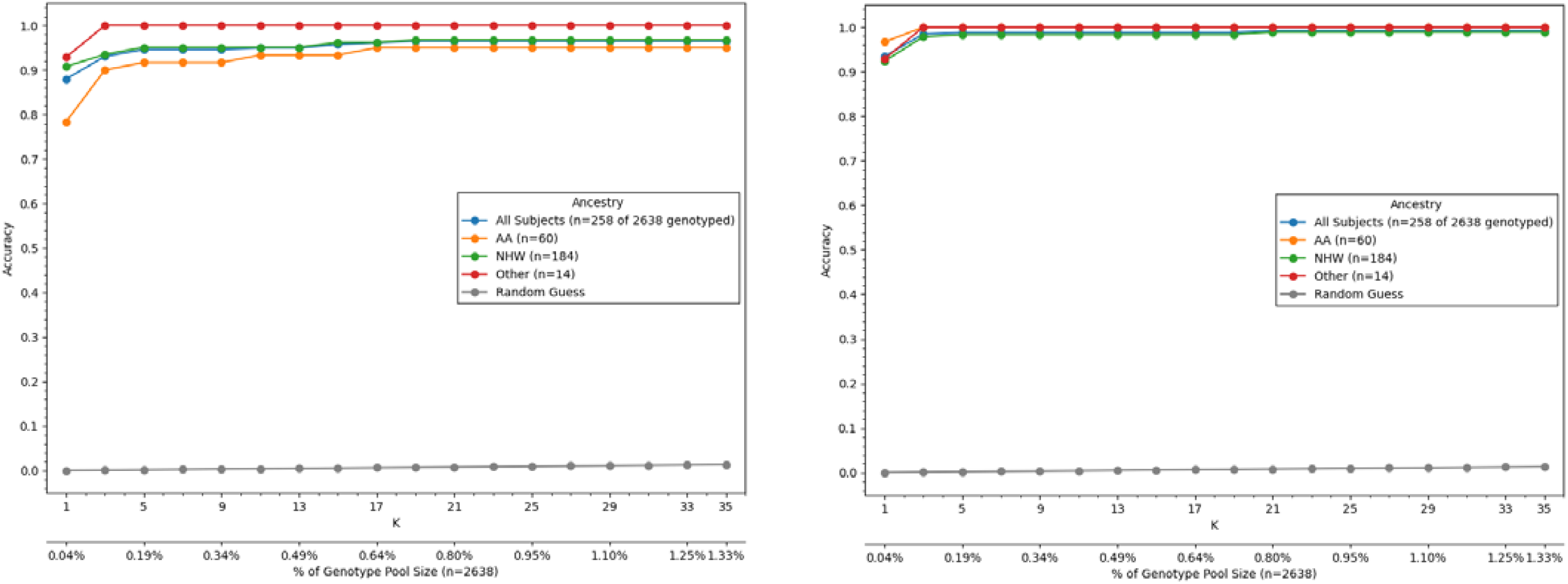
Training with data from diverse populations improves testing accuracy in African Americans (AA). (A) First attempts at training with only 13% AA subjects in SPIROMICS resulted in lower testing accuracy in independent AA compared to non-Hispanic White (NHW) subjects. (B) After training with both COPDGene and JHS subjects, identification accuracy significantly improved in AA subjects, but was still slightly lower than NHW subjects.

Next, we sought to determine the minimum number of protein-pQTL pairs that were necessary to match a proteome to a genome. First, we ranked protein-pQTL pairs by p-value and then retested using only smaller subsets of the strongest protein-pQTL pairs (**Supplemental Table 1**). In general, overall accuracy plateaued at around 100 protein-pQTLs, suggesting that this was the minimum number of proteins measurements that were needed to confidently identify a subject’s genotype using only protein data.

### Testing accuracy of matching proteome to genome across diverse, independent cohorts

Using the top 100 protein-pQTL SNPs from the training data using (COPDGene and JHS training subjects), we then tested prediction accuracies in 4 cohorts (SPIROMICS, MESA, JHS, COPDGene) using independent subjects that had not been used for training, including accuracies based on race and ethnicity (**Table 2**). The true match was among the highest odds for most subjects (>85%) in the cohorts and populations, except for COPDGene and Black Americans in MESA. If we took the top 1% of highest odds, the true match was among the highest odds for most subjects (>85%) in all cohorts and populations.

To determine whether newer and larger proteome assays were more or less accurate at identifying genetic profiles, we randomly split 5,292 COPDGene subjects who had SomaScan v4.0 5k data (4,776 proteins) into training and testing groups using a 50/50 train-test split (**Supplemental Table 2**) to generate a new list of protein-pQTL pairs (**Supplemental File 2**). With as few as 100 proteins, identification accuracy improved to >99% (Table 4) and accuracy in subjects with African ancestry was similar to those with predominantly European ancestry (**Supplemental Table 3**).

**Table 4:** Optimizing of number of training proteins for SomaScan 5K dataset

SD – standard deviation;
|  |  |  |  |  |  |  |
| --- | --- | --- | --- | --- | --- | --- |
|  | COPDGene Training |  |  | COPDGene Testing |  |  |
|  | % correctly identified subjects |  |  | % correctly identified subjects |  |  |
| # proteins | Top 1 | In top 3 | In top 1% | Top 1 | In top 3 | In top 1% |
| 20 | 78.34% | 88.55% | 98.41% | 76.91% | 87.76% | 98.34% |
| 40 | 97.85% | 99.06% | 99.77% | 96.98% | 98.56% | 99.66% |
| 60 | 99.06% | 99.51% | 99.77% | 98.30% | 99.17% | 99.66% |
| 100 | 99.51% | 99.66% | 99.77% | 99.09% | 99.43% | 99.66% |
| 150 | 99.62% | 99.70% | 99.77% | 99.28% | 99.58% | 99.70% |
| 250 | 99.70% | 99.74% | 99.77% | 99.36% | 99.62% | 99.66% |
| All | 97.01% | 97.54% | 98.90% | 92.44% | 94.29% | 97.43% |

### Genome privacy protection through proteome transformation

Since we have shown that measurement of selected proteins with strong pQTLs represent genetic equivalents, we reasoned that removing the pQTL effects on the proteome would inhibit the ability to reidentify a subject. One method that accomplishes this is to adjust each protein measurement by subtracting the population mean for that genotype (**Figure 6**). This method has the advantage in that if the subject’s genotype and the correction factors are known, it is simple to recapitulate the actual protein measurements. In both testing cohorts, subtracting the genotype effect abolished the ability to identify subjects (**Figure 7)**.

**Figure 6.**
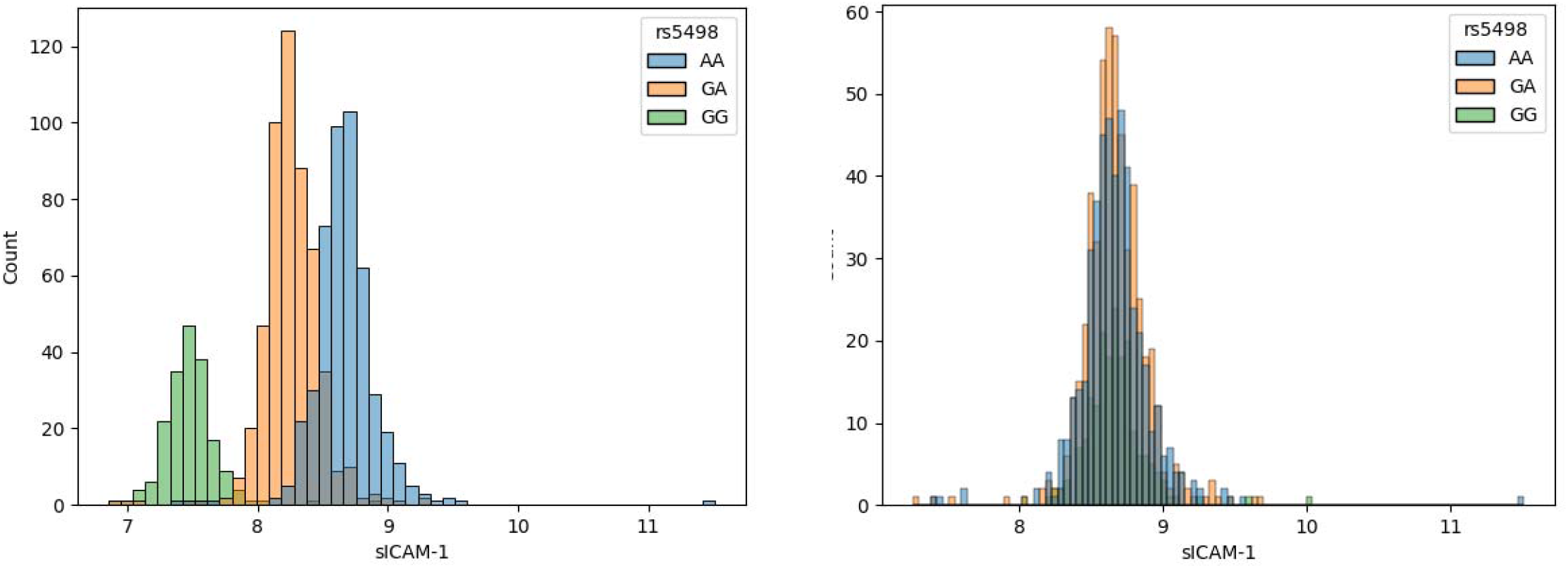
Poisoning data by adjusting protein values for genotype. (A). sICAM histograms showing normal probability distribution functions for sICAM-1, which have been log transformed. In this example AA is the major genotype. (B). Adjusting protein levels by recentering the mean on each genotype group abolishes the genotype effect on sICAM-1 measurements.

**Figure 7.**
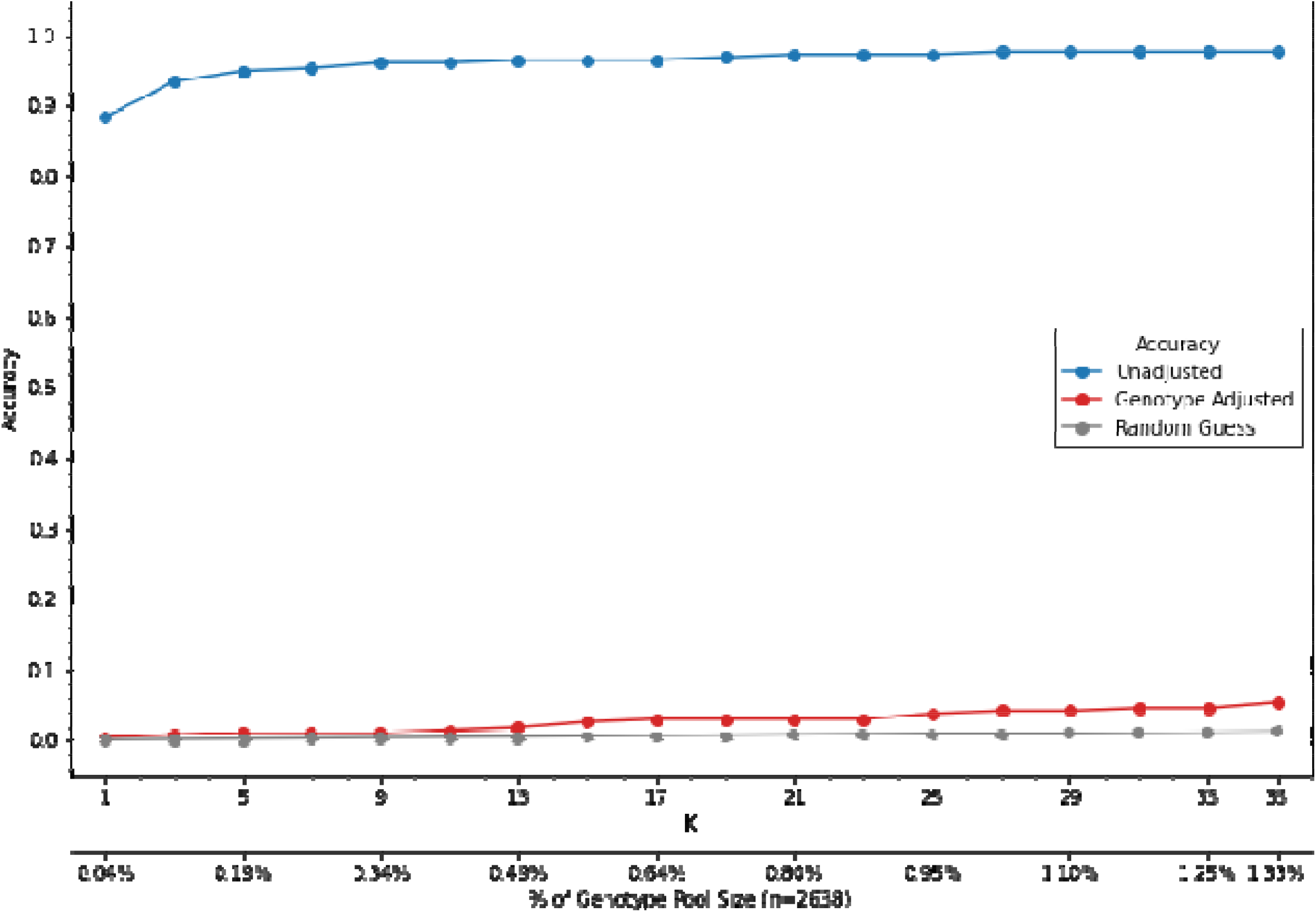
Removing the mean protein-pQTL effect abolishes the ability of matching a proteome to a genome. Shown are accuracy of matching algorithm with (red) and without (blue) removing mean pQTL effect as well as the probably of a random guess matching (grey).

### Can genotype adjustment preserve biomarker-phenotype associations?

To test if adjusting for genotype affects associations between biomarkers and phenotypes, we first identified two proteins, sICAM-5 and DERM, which were significantly associated with smoking status in both the COPDGene and SPIROMICS testing cohorts. Next, we assessed the association before and after adjustment for genotype. In both cohorts, associations with smoking status did not change significantly after genotype adjustment (**Supplemental Table 4**).

## Discussion

De-identification of data is a key concept for shared research and privacy protection but is not yet used in large scale proteomic studies. While proof of concept studies have suggested that specific amino acid peptides with missense variants (minor allelic peptides) can identify genotype variants in some subjects [15], this approach requires mass spectrometry and has not yet been used across large scale cohort studies with proteomic data. This study is the first to demonstrate on a large scale that proteomic data are not privacy protected because an individual proteome can be matched to a specific genome with high accuracy even without protein sequence information. The key identifying features in the proteome are the effects of common pQTLs, which link a measured protein level to a specific genotype. Furthermore, we show that identification only requires a small number of proteins (as few as 60-100 selected proteins) to link an individual protein profile to a single genetic profile among thousands of subjects. Additionally, our results suggest that using diverse subjects for selecting the most influential proteins improves overall accuracy, particularly among those with African ancestry and underscores the importance of including diverse subjects in Omics research. Finally we show that proteomic data can identify behavioral features (e.g., smoking). The ability to accurately infer genetic ancestry and also characterize behavioral features implies that proteomic data should have the at least the same (if not more rigorous) privacy protections as genomic datasets.

The two main technological breakthroughs that have facilitated accurately matching an individual proteome to a specific genome are improvement in high throughput proteomic technologies and large scale pQTL studies. Until the last few years, there were no proteomic platforms that could simultaneously and accurately measure more than 100 proteins and there was little known about which of those proteins had strong pQTLs. While our study used two different SomaScan platforms, lack of privacy (de-identification) should be implied for any platform that can simultaneously measure thousands of proteins even when mass spectrometry is not used. The logical continuation of this principle is that proteomic data could be used to discriminate based on identifying the sex of a subject, ancestry, or paternity. A protein profile could even be used to identify close relatives for forensic purposes.

De-identification and privacy protection by informatics is a growing field. While we demonstrate that the simple method of removing the pQTL effect of proteins can significantly degrade the ability to link a proteome to a genome, we recognize the large body of emerging literature on alternative data obfuscation methods to protect privacy of many types of data [16]. These methods range from industry level data obfuscation/masking and secure data outsourcing techniques such as substitution, shuffling, numeric variance and null-out/mask-out, to more rigorous statistical data obfuscation methodologies used in Hippocratic Databases [17], and privacy-preserving data mining [18] such as k-anonymization [19], l-diversity [20], t-Closeness [21], differential-privacy [22] based methods. Machine learning [Barla, 2008 #44] and deep learning [Wen, 2020 #45] are also being used in proteomic feature identification and we may be able to leverage these same methods to isolate and “cloak” identifiable omics features while maintaining desirable statistical properties of the data for downstream application.

Bioethicists had anticipated that other omics data such as proteomic data might one day be identifiable and create privacy concerns [23] and our work demonstrates that this day has come even for proteomic technologies that do not rely on peptide sequencing. Unfortunately, most governmental policies do not yet apply to newer omics data such as proteomics (one exception may be the General Data Protection Regulation in the European Union, which protects biological equivalents of genotypes). We suggest biomedical research policies be clarified or amended to include any omics data (e.g., measurement of proteins or other molecules, such as metabolites) in which genotype can be ascertained [24], but also that there be consideration beyond genotype equivalents to include all features of omics (e.g. behavioral information such as smoking). Because data protection is imperfect and frequently breached, a complementary solution to maintaining privacy might include bioinformatic and cryptographic adjustments to proteomic data. We demonstrated that adjusting out the genetic effects on protein measurements protects privacy by obfuscating the genetic effects, but it still does not change non-genetic associations (such as smoking). This strategy is simple and can be reversed if necessary, when a researcher has the accompanying genetic information. A disadvantage to removing genetic coding of the proteome is that it could remove associations in which genotype mediates protein affect. Another caveat from our work is that if training the method does not include diverse populations, the methods may not be generalizable outside European ancestry. Thus, it is important that future genomic and proteomic work include more underrepresented populations.

## Data Availability

All data produced in the present study are available upon reasonable request to the authors.

## Author Contributions

**Conceived and designed manuscript:** RPB AH

**Performed the Experiments:** RPB CCQ NR DJ KU GM

**Analyzed the Data**: AH, EML

**Contributed reagents/materials/analysis tools:** AH, EML

**Wrote the Paper:** RPB KK LG AH

**Critically reviewed the Paper:** all authors

## Funding Support and Acknowledgements

### COPDGene

The project described was supported by Award Number U01 HL089897 and Award Number U01 HL089856 from the National Heart, Lung, and Blood Institute. The content is solely the responsibility of the authors and does not necessarily represent the official views of the National Heart, Lung, and Blood Institute or the National Institutes of Health.

COPD Foundation Funding

COPDGene is also supported by the COPD Foundation through contributions made to an Industry Advisory Board comprised of AstraZeneca, Boehringer-Ingelheim, Genentech, GlaxoSmithKline, Novartis, Pfizer, Siemens, and Sunovion.

COPDGene® Investigators – Core Units

*Administrative Center*: James D. Crapo, MD (PI); Edwin K. Silverman, MD, PhD (PI); Barry J. Make, MD; Elizabeth A. Regan, MD, PhD

*Genetic Analysis Center*: Terri Beaty, PhD; Ferdouse Begum, PhD; Peter J. Castaldi, MD, MSc; Michael Cho, MD; Dawn L. DeMeo, MD, MPH; Adel R. Boueiz, MD; Marilyn G. Foreman, MD, MS; Eitan Halper-Stromberg; Lystra P. Hayden, MD, MMSc; Craig P. Hersh, MD, MPH; Jacqueline Hetmanski, MS, MPH; Brian D. Hobbs, MD; John E. Hokanson, MPH, PhD; Nan Laird, PhD; Christoph Lange, PhD; Sharon M. Lutz, PhD; Merry-Lynn McDonald, PhD; Margaret M. Parker, PhD; Dmitry Prokopenko, Ph.D; Dandi Qiao, PhD; Elizabeth A. Regan, MD, PhD; Phuwanat Sakornsakolpat, MD; Edwin K. Silverman, MD, PhD; Emily S. Wan, MD; Sungho Won, PhD

*Imaging Center*: Juan Pablo Centeno; Jean-Paul Charbonnier, PhD; Harvey O. Coxson, PhD; Craig J. Galban, PhD; MeiLan K. Han, MD, MS; Eric A. Hoffman, Stephen Humphries, PhD; Francine L. Jacobson, MD, MPH; Philip F. Judy, PhD; Ella A. Kazerooni, MD; Alex Kluiber; David A. Lynch, MB; Pietro Nardelli, PhD; John D. Newell, Jr., MD; Aleena Notary; Andrea Oh, MD; Elizabeth A. Regan, MD, PhD; James C. Ross, PhD; Raul San Jose Estepar, PhD; Joyce Schroeder, MD; Jered Sieren; Berend C. Stoel, PhD; Juerg Tschirren, PhD; Edwin Van Beek, MD, PhD; Bram van Ginneken, PhD; Eva van Rikxoort, PhD; Gonzalo Vegas Sanchez-Ferrero, PhD; Lucas Veitel; George R. Washko, MD; Carla G. Wilson, MS;

*PFT QA Center, Salt Lake City, UT*: Robert Jensen, PhD

*Data Coordinating Center and Biostatistics, National Jewish Health, Denver, CO*: Douglas Everett, PhD; Jim Crooks, PhD; Katherine Pratte, PhD; Matt Strand, PhD; Carla G. Wilson, MS

*Epidemiology Core, University of Colorado Anschutz Medical Campus, Aurora, CO*: John E. Hokanson, MPH, PhD; Gregory Kinney, MPH, PhD; Sharon M. Lutz, PhD; Kendra A. Young, PhD

*Mortality Adjudication Core:* Surya P. Bhatt, MD; Jessica Bon, MD; Alejandro A. Diaz, MD, MPH; MeiLan K. Han, MD, MS; Barry Make, MD; Susan Murray, ScD; Elizabeth Regan, MD; Xavier Soler, MD; Carla G. Wilson, MS

*Biomarker Core*: Russell P. Bowler, MD, PhD; Katerina Kechris, PhD; Farnoush Banaei-Kashani, Ph.D

BDH is supported by NIH K08 HL136928, U01 HL089856, R01 HL135142, R01 HL139634, and R01 HL147148.

## SPIROMICS Acknowledgement and Funding Statement

The authors thank the SPIROMICS participants and participating physicians, investigators and staff for making this research possible. More information about the study and how to access SPIROMICS data is available at www.spiromics.org. The authors would like to acknowledge the University of North Carolina at Chapel Hill BioSpecimen Processing Facility for sample processing, storage, and sample disbursements (http://bsp.web.unc.edu/). We would like to acknowledge the following current and former investigators of the SPIROMICS sites and reading centers: Neil E Alexis, MD; Wayne H Anderson, PhD; Mehrdad Arjomandi, MD; Igor Barjaktarevic, MD, PhD; R Graham Barr, MD, DrPH; Patricia Basta, PhD; Lori A Bateman, MSc; Surya P Bhatt, MD; Eugene R Bleecker, MD; Richard C Boucher, MD; Russell P Bowler, MD, PhD; Stephanie A Christenson, MD; Alejandro P Comellas, MD; Christopher B Cooper, MD, PhD; David J Couper, PhD; Gerard J Criner, MD; Ronald G Crystal, MD; Jeffrey L Curtis, MD; Claire M Doerschuk, MD; Mark T Dransfield, MD; Brad Drummond, MD; Christine M Freeman, PhD; Craig Galban, PhD; MeiLan K Han, MD, MS; Nadia N Hansel, MD, MPH; Annette T Hastie, PhD; Eric A Hoffman, PhD; Yvonne Huang, MD; Robert J Kaner, MD; Richard E Kanner, MD; Eric C Kleerup, MD; Jerry A Krishnan, MD, PhD; Lisa M LaVange, PhD; Stephen C Lazarus, MD; Fernando J Martinez, MD, MS; Deborah A Meyers, PhD; Wendy C Moore, MD; John D Newell Jr, MD; Robert Paine, III, MD; Laura Paulin, MD, MHS; Stephen P Peters, MD, PhD; Cheryl Pirozzi, MD; Nirupama Putcha, MD, MHS; Elizabeth C Oelsner, MD, MPH; Wanda K O’Neal, PhD; Victor E Ortega, MD, PhD; Sanjeev Raman, MBBS, MD; Stephen I. Rennard, MD; Donald P Tashkin, MD; J Michael Wells, MD; Robert A Wise, MD; and Prescott G Woodruff, MD, MPH. The project officers from the Lung Division of the National Heart, Lung, and Blood Institute were Lisa Postow, PhD, and Lisa Viviano, BSN; SPIROMICS was supported by contracts from the NIH/NHLBI (HHSN268200900013C, HHSN268200900014C, HHSN268200900015C, HHSN268200900016C, HHSN268200900017C, HHSN268200900018C, HHSN268200900019C, HHSN268200900020C), grants from the NIH/NHLBI (U01 HL137880 and U24 HL141762), and supplemented by contributions made through the Foundation for the NIH and the COPD Foundation from AstraZeneca/MedImmune; Bayer; Bellerophon Therapeutics; Boehringer-Ingelheim Pharmaceuticals, Inc.; Chiesi Farmaceutici S.p.A.; Forest Research Institute, Inc.; GlaxoSmithKline; Grifols Therapeutics, Inc.; Ikaria, Inc.; Novartis Pharmaceuticals Corporation; Nycomed GmbH; ProterixBio; Regeneron Pharmaceuticals, Inc.; Sanofi; Sunovion; Takeda Pharmaceutical Company; and Theravance Biopharma and Mylan.

## MESA acknowledgement

The MESA project is supported by the National Heart, Lung, and Blood Institute (NHLBI) in collaboration with MESA investigators. Support for MESA is provided by contracts 75N92020D00001, HHSN268201500003I, N01-HC-95159, 75N92020D00005, N01-HC-95160, 75N92020D00002, N01-HC-95161, 75N92020D00003, N01-HC-95162, 75N92020D00006, N01-HC-95163, 75N92020D00004, N01-HC-95164, 75N92020D00007, N01-HC-95165, N01-HC-95166, N01-HC-95167, N01-HC-95168, N01-HC-95169, UL1-TR-000040, UL1-TR-001079, and UL1-TR-001420. Also supported in part by the National Center for Advancing Translational Sciences, CTSI grant UL1TR001881, and the National Institute of Diabetes and Digestive and Kidney Disease Diabetes Research Center (DRC) grant DK063491 to the Southern California Diabetes Endocrinology Research Center. Infrastructure for the CHARGE Consortium is supported in part by the National Heart, Lung, and Blood Institute (NHLBI) grant R01HL105756.

Molecular data for the Trans-Omics in Precision Medicine (TOPMed) program was supported by the National Heart, Lung and Blood Institute (NHLBI). SOMAscan proteomics for NHLBI TOPMed: Multi-Ethnic Study of Atherosclerosis (MESA)” (phs001416.v1.p1) was performed at the Broad Institute and Beth Israel Proteomics Platform (HHSN268201600034I). Core support including centralized genomic read mapping and genotype calling, along with variant quality metrics and filtering were provided by the TOPMed Informatics Research Center (3R01HL-117626-02S1; contract HHSN268201800002I). Core support including phenotype harmonization, data management, sample-identity QC, and general program coordination were provided by the TOPMed Data Coordinating Center (R01HL-120393; U01HL-120393; contract HHSN268201800001I). We gratefully acknowledge the studies and participants who provided biological samples and data for TOPMed.

### JHS acknowledgement

The Jackson Heart Study (JHS) is supported and conducted in collaboration with Jackson State University (HHSN268201800013I), Tougaloo College (HHSN268201800014I), the Mississippi State Department of Health (HHSN268201800015I) and the University of Mississippi Medical Center (HHSN268201800010I, HHSN268201800011I and HHSN268201800012I) contracts from the National Heart, Lung, and Blood Institute (NHLBI) and the National Institute for Minority Health and Health Disparities (NIMHD). The authors also wish to thank the staffs and participants of the JHS.

## Disclaimers

The views expressed in this manuscript are those of the authors and do not necessarily represent the views of the National Heart, Lung, and Blood Institute; the National Institutes of Health; or the U.S. Department of Health and Human Services

---

**Supplemental File 1 – List of training proteins and SNP**

See online supplement

**Supplemental Table 1:** Optimizing of number of training proteins

| **Supplemental Table 1: Optimizing of number of training proteins** |  |  |  |  |  |  |
| --- | --- | --- | --- | --- | --- | --- |
|  | COPDGene Training |  |  | JHS Training |  |  |
|  | % correctly identified subjects |  |  | % correctly identified subjects |  |  |
| # proteins | Top 1 | In top 3 | In top 1% | Top 1 | In top 3 | In top 1% |
| 20 | 68.7% | 79.8% | 95.3% | 53.8% | 73.2% | 93.6% |
| 40 | 82.5% | 89.7% | 96.8% | 79.9% | 89.0% | 97.8% |
| 60 | 88.1% | 93.2% | 97.5% | 85.7% | 92.3% | 98.4% |
| 100 | 92.1% | 94.8% | 97.7% | 91.1% | 95.1% | 99.3% |
| 150 | 94.3% | 96.5% | 98.1% | 93.2% | 96.7% | 99.6% |
| 250 | 93.0% | 95.1% | 97.8% | 92.5% | 96.8% | 99.7% |
| All | 91.0% | 93.3% | 96.2% | 92.3% | 96.4% | 99.6% |

**Supplemental Table 2:** Characteristics of training cohort and independent testing cohorts with SomaScan 5k

|  | Training | Testing |
| --- | --- | --- |
| Cohort | COPDGene<br>(N = 2,646 with SomaScan<br>N = 2,646 genotyped) | COPDGene<br>(N = 2,646 with SomaScan<br>N = 9,970 genotyped) |
| Gender (%female) | 50.6% | 48.4% |
| Age (± SD) | 65.3 ± 8.5 | 65.7 ± 8.7 |
| Race/ethnicity (self-reported) |  |  |
| White, non-Hispanic | 70.9% | 70.7% |
| Black, non-Hispanic | 29.1% | 29.3% |

SD – standard deviation;

**Supplemental Table 3.** Accuracy of matching proteome profiles to genetic profiles using SomaScan 5k

|  |  | % correctly identified |  |  |
| --- | --- | --- | --- | --- |
| Testing Cohort | Subgroup | Top 1 | In top 3 | In Top 1 % |
| COPDGene | Overall | 99.28% | 99.58% | 99.70% |
|  | NHW | 99.47% | 99.63% | 99.63% |
|  | AFA | 98.84% | 99.48% | 99.87% |

**Supplemental Table 4:** Association with smoking status is preserved after adjusting for genetic effect.

|  |  |  |  |  |  |  |  |
| --- | --- | --- | --- | --- | --- | --- | --- |
|  |  | No Genotype Adjustment |  |  | Genotype Adjustment |  |  |
| **Dataset** | **Protein** | **t-statistic** | **smoking difference** | **p** | **t-statistic** | **smoking difference** | **p** |
| SPIROMICS | DERM | 5.64 | 0.181 | 6.0E-08 | 5.54 | 0.176 | 9.4E-08 |
|  | sICAM-5 | -3.31 | -0.150 | 0.0012 | -3.33 | -0.137 | 0.0011 |
| COPDGene | DERM | 9.86 | 0.139 | 5.5E-22 | 9.63 | 0.133 | 4.7E-21 |
|  | sICAM-5 | -4.70 | -0.087 | 3.0E-06 | -5.45 | -0.092 | 6.5E-08 |

DERM – dermatopontin; sICAM-5: soluble Intracellular adhesion molecule 5; smoking difference the mean of current smokers – mean never and former smokers

**Supplemental File 2** –pQTL SNPs from SomaScan 5k Discovery

See online supplement

